# Mapping Referred Sensation In Traumatic Brachial Plexus Injury Patients

**DOI:** 10.1101/2024.02.19.23297564

**Authors:** Ana Carolina Schmaedeke, Fátima S. Erthal, Claudia D. Vargas, Bia L. Ramalho

## Abstract

**Background:** Referred sensation (RS) is described as the sensation evoked in the skin areas other than the stimulated region. The aim of the study was to identify and map RS in traumatic brachial plexus injury (TBPI) patients through a standardised assessment.

**Methods:** In Experiment 1, 12 patients underwent an RS screening by stimulating 22 skin areas distributed in both upper limbs, neck, and face with a cotton swab. In Experiment 2, a detailed RS mapping employing Semmes-Weinstein monofilament was performed at the reinnervated forearm of three subjects who showed RS in experiment 1 screening. In one of these patients, RS mapping was performed over a time span of 6.3 years.

**Results:** Screening of TBPI patients submitted to different reconstructive surgeries revealed RS only in the patients who underwent intercostal to musculocutaneous nerve (ICN-MCN) transfer. RS systematic mapping of these patients revealed a unique distribution in the forearm and the chest, without any clear topographic organisation. Longitudinal assessment in one of the tested participants showed a scattered expansion of the RS throughout time.

**Conclusions:** This is the first study to map systematically the RS in patients with TBPI. RS was identified only after ICN-MCN transfer. A possible explanation for this phenomenon is that the greater distance between the representations of the donor and receptor nerves at the primary somatosensory cortex (S1) could constrain plastic reorganisation and thus limit the disentangling of the forearm and chest sensations.

## Introduction

Referred sensation (RS) is described as the sensation evoked by stimulating the skin surface in a place other than that of the stimulated region ^1^. Although RS has been reported in able-body individuals ^2,3^, it is commonly described after severe peripheral injury ^4,5^. Cortical reorganisation and disorganised axonal growth are considered as putative conditions for the RS appearance after a peripheral injury ^6,7^. Cortical sensorimotor regions often respond to adjacent territories after the loss of their primary inputs (e.g., invasion of the face representation into the cortical area of the hand after hand amputation), possibly through the unmasking of silent synapses ^7–12^. This reorganisation can also be due to increased inputs from other intact body parts ^13–15^. Alternatively, the remaining axons can grow towards targets distinct from those they originally innervated ^6,16,17^.

Besides amputation, the brachial plexus injury is also described as causing referred sensation ^18–21^. The brachial plexus is the structure formed by five nerve roots (C5, C6, C7, C8 and T1) responsible for the motor, sensory and autonomic innervation of the upper limb ^22,23^. The motor deficit resulting from TBPI varies according to the injured roots and can lead to significant functional decline of the upper limb ^24,25^. Along with motor damage, there is an loss or decrease in sensitivity in the affected limb, often associated with pain as well as referred and phantom sensations ^18–21^.

TBPI treatments available today consist of physiotherapy and surgeries aiming at the reinnervation of the segment affected by the injury. A wide variety of surgical techniques can be combined according to the extent of the injury, usually by taking the motor deficits into consideration ^26,27^. A surgical procedure performed quite often after TBPI is nerve transfer, where an intact donor nerve or fascicle is transferred to the injured nerve as distally as possible to reinnervate an injury-affected muscle ^24,28^. In this procedure, special care is taken to isolate the motor component of the donor nerve so as to provide motor recovery through the reinnervation of the recipient muscle ^29^. In this context lies the anecdotal description of RS in TBPI ^22,30,31^.

The presence of RS after nerve transfer in TBPI raises the question of whether or not these surgically-induced new sensory connections can be integrated into the body map representation through cortical reorganisation. As a first step, surgically-induced RS after a TBPI must be accessed in detail. Therefore, the aim of this study was to: 1) identify RS in TBPI patients that underwent different surgical procedures through a standardised assessment and 2) map surgically-induced RS in patients who underwent intercostal to musculocutaneous nerve (ICNs-MCN) transfer through a longitudinal follow-up.

## Material and methods

### Participants

Thirteen TBPI patients were recruited from the physical therapy facility of the Institute of Neurology Deolindo Couto (INDC-UFRJ). These subjects were clinically evaluated and registered in the laboratory’s digital database on TBPI ^32^, and data were stored on the Neuroscience Experiment System (NES) platform ^33^.

Inclusion criteria were age equal to 18 years or older, preserved communication skills, and unilateral TBPI diagnosed through clinical and complementary exams. Exclusion criteria were previous primary or secondary central and/or peripheral nervous system diseases. All participants provided written informed consent to participate in the study, which was in accordance with the declaration of Helsinki and has been approved by the ethics committee of the Institute of Neurology Deolindo Couto-UFRJ, Brazil (process number: 298.925).

## Sensory evaluation

### Experiment 1: Referred sensation screening

Twenty-two stimulation points distributed between both upper limbs, neck, and face were selected for a tactile screening to identify the referred sensations’ location (Figure 1.A). These points corresponded to the dermatomes of the roots from C2 to T2 and to the lower branches of the trigeminal nerve. Stimulation was performed with a cotton swab while subjects were comfortably seated and blindfolded. The order of stimulation was pseudo-randomized. Each point was stimulated three times consecutively and participants were then asked to report if they felt, what they felt (based on a list of sensations but no limited to - pricking, burning, squeeze, pressure, vibration, movement, electric shock, touch, itch, heat, cold, tickle, tingling and sweat (free translation from Portuguese)) as well as the location of the sensation, and this information was recorded on a paper sheet. Subjects could point out the sensation site with the uninjured upper limb. If the location of the evoked sensation did not correspond to the stimulated point, it was considered a referred sensation (RS). If, during the stimuli, no sensation was evoked at a particular point, then it was considered a point of anaesthesia. To avoid any possible bias demand characteristics of the experimental protocol^34^, we have decided to not indicate to the participants that the sensation could be felt in more than one place.

**Figure 1.**
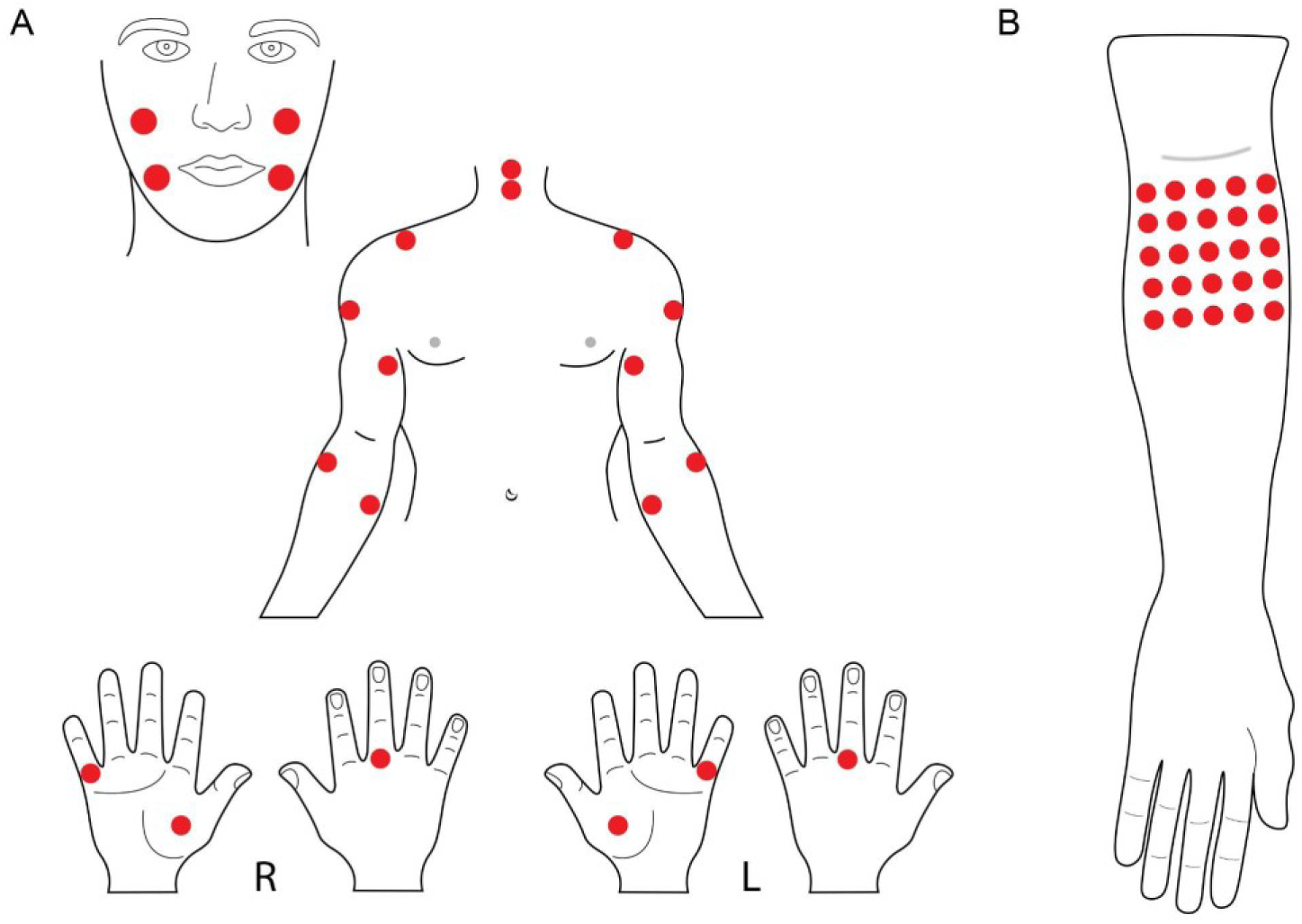
Representation of the stimulated points. **(A)** referred sensation screening in experiment 1. R: right hand, L: left hand; **(B)** mapping of the referred sensations in the forearm associated to the intercostal to musculocutaneous nerve transfer in experiment 2.

Thus, before the beginning of the experimental trial the participant was instructed as follows:

> *“Now an evaluation will be made with a cotton swab. This evaluation will be carried out with yourself blindfolded. In this case, I will touch some regions of your body with the cotton swab and you must tell me what you are feeling, that means, “how does the sensation feel?” - you must answer with, for example, pricking, burning, squeeze, pressure, vibration, movement, electric shock, touch, itch, heat, cold, tickle, tingling and sweat. After this, you must tell me exactly where you are feeling it (any region of your body). You should be comfortably seated and must remain focused on the assessment at all times.”*

### Experiment 2: Sensorial mapping of the referred sensation

During the screening performed in experiment 1, two participants (BPI11 and BPI12) consistently reported evoked sensations in the chest when the lateral forearm was stimulated. These two subjects had undergone ICN-MCN transfer. In order to map the chest referred sensations, a new protocol designed for this purpose was applied. The protocol was adapted from that used in amputees to map the sensation of the amputated limb after surgical targeted sensory reinnervation of a pectoral region ^35,36^. Thus, a grid of points was drawn with a dermographic pencil around the autonomous zone of the musculocutaneous nerve ^37^ in the patient’s injured forearm (Figure 1.B) to standardise the stimulation area. For the stimulation, the filament with the greatest calibre from a set of 20 Semmes-Weinstein Monofilaments (SWM, Bioseb, Vitrolles, France) was chosen to ensure that participants felt the stimuli, avoiding false negatives (based on our previous findings of increased touch thresholds in TBPI patients - ^21^. Furthermore, with this filament it was possible to stimulate in restricted skin spots, allowing a detailed mapping of the forearm. This filament exerts a force of 160 grams (or 6.20 log of the force (10 x force in mg)) when applied at 90° over the skin, bending into a “C” shape (Ramalho et al., 2019). Participants were asked to sit comfortably, with bare chest, blindfolded, with the injured arm resting on a pillow. The grid points were then stimulated by applying the filament once at each point. The patients were instructed to say “yes” every time they felt something, then the evaluator asked them to report the type of the evoked sensation and to point out with the uninjured upper limb the region where the sensation was felt. When the referred sensation was reported, its location was delimited with a dermographic pencil.

The following instruction was given before experiment 2:

> *“I will start the evaluation by touching a region of the grid on your right (or left) arm with this filament and you should say “Yes” to me each time you feel the touch. Then you must tell me what you felt and point out where you felt it in your body. You can feel it in more than one place at the same time, for example, in two places on your arm or on your arm and chest. After indicating the location of the sensation, I will mark it with the pencil I used to make the dot grid.”*

One subject (BPI13), who was not assessed at the referred sensation screening (experiment 1), was included in experiment 2. This subject had undergone ICN-MCN transfer and participated in a touch threshold assessment of both arms in Ramalho et al., (2019) (see Supplementary Figure 2, participant BPI16 - Ramalho et al. 2019). He was the only subject evaluated longitudinally.

## Results

### Participants

Thirteen BPI participants were investigated (2 females; mean age: 32 years ± 6.22, range: 19-40). Personal information about the participants, the extent of the injury, type of surgery, time between injury and surgery and time between injury and sensory assessment can be found in Table 1. Six patients had all brachial plexus roots compromised (C5-T1), and 7 patients had lesions in the upper trunk (C5-C6) or extended upper trunk (C5-C7). One participant had not undergone any surgery at the time of the evaluation, while two patients underwent two surgeries. The interval between the injury and the first surgery ranged from 86 to 372 days (mean: 184 days ± 97,94), while the interval between the injury and the sensory assessment ranged from 165 to 1396 days (mean: 603 ± 405.95).

**Table 1:** Subjects characteristics, description of brachial plexus lesion ⁄ surgery.

Time periods are expressed in days.
| ID | Gender, Age Range (years) | Handedness | Injury side and extension | Time between injury and surgery | Surgery type | Time between surgery and screening | Time between surgery and mapping | Referred sensation |
| --- | --- | --- | --- | --- | --- | --- | --- | --- |
| BPI01 | F, 26-30 | right-handed | Left, C5-T1 | 195 | SAN-SSN transfer | 857 | - | No |
| BPI02 | M, 26-30 | right-handed | Left, C5-T1 | 372 | Exploration + C5, C6, C7 and SSN neurolysis | 81 | - | No |
| BPI03 | M, 26-30 | right-handed | Right, C5-T1 | 185 | Exploration | 284 | - | No |
| BPI04 | M, 31-35 | right-handed | Left, C5-C7 | 141 | Oberlin nerve transfer | 28 | - | No |
| BPI05 | M, 36-40 | right-handed | Left, C5-C7 | 355 | Oberlin nerve transfer | 45 | - | No |
| BPI06 | M, 31-35 | right-handed | Right, C5-C7 | 148 | SAN-SSN transfer + UN-MCN transfer | 63 | - | No |
| BPI07 | F, 26-30 | right-handed | Right, C5-C7 | 260 | SAN-MCN transfer / Oberlin nerve transfer | 871 | - | No |
| BPI08 | M, 21-25 | right-handed | Right, C5-C7 | - | no surgery | 534** | - | No |
| BPI09 | M, 31-35 | right-handed | Right, C5-C6 | 177 | SAN-SSN transfer + Oberlin nerve transfer + TRN-AXN transfer | 1219 | - | No |
| BPI10 | M, 18-20 | right-handed | Right, C5-C7 | 100 | Oberlin nerve transfer | 126 | - | No |
| BPI11 | M, 31-35 | ambidextrous | Left, C5-T1 | 92 | SAN-SSN transfer + ICNs-MCN transfer | 644 | 707 | **Yes** |
| BPI12 | M, 36-40 | ambidextrous | Left, C5-T1 | 102 | ICNs-MCN transfer / SSN neurolysis | 798 | 833 | **Yes** |
| BPI13 | M, 36-40 | right-handed | Left, C5-T1 | 86 | SAN-SSN transfer + ICNs-MCN transfer | - | 79<br>758,<br>2242 | No<br>**Yes** |

In patients with a complete brachial plexus injury (C5-T1), the transfer of the spinal accessory nerve to the suprascapular nerve (SAN-SSN) and the transfer of INCs-MCN were the most common surgical interventions. In patients with upper trunk lesions, the transfer of a fascicle of the ulnar nerve to the biceps motor branch of MCN (Oberlin nerve transfer) was the most common.

## Sensory evaluation

### Experiment 1: Referred sensation (RS) screening

The results of the RS screening can be seen in Figure 2. Each picture represents a patient’s sensory deficit profile. Nine subjects did not feel the stimulation at one or more stimulated points, hereafter considered as points of anaesthesia (black crosses). BPI01 had the worst sensory deficit, as none of the stimulated points on the injured arm evoked any sensation, while BPI09 and BPI10 had no identified anaesthesia points. Two participants (BPI11 and BPI12) consistently reported RS (orange circles), while the remaining participants submitted to this evaluation did not report any RS. BPI11 presented anaesthesia when the left hand points and the medial point of the left forearm were stimulated. Both BPI11 and BPI 12 reported RS in the chest when the lateral point of the forearm was stimulated (Fig. 2).

**Figure 2.**
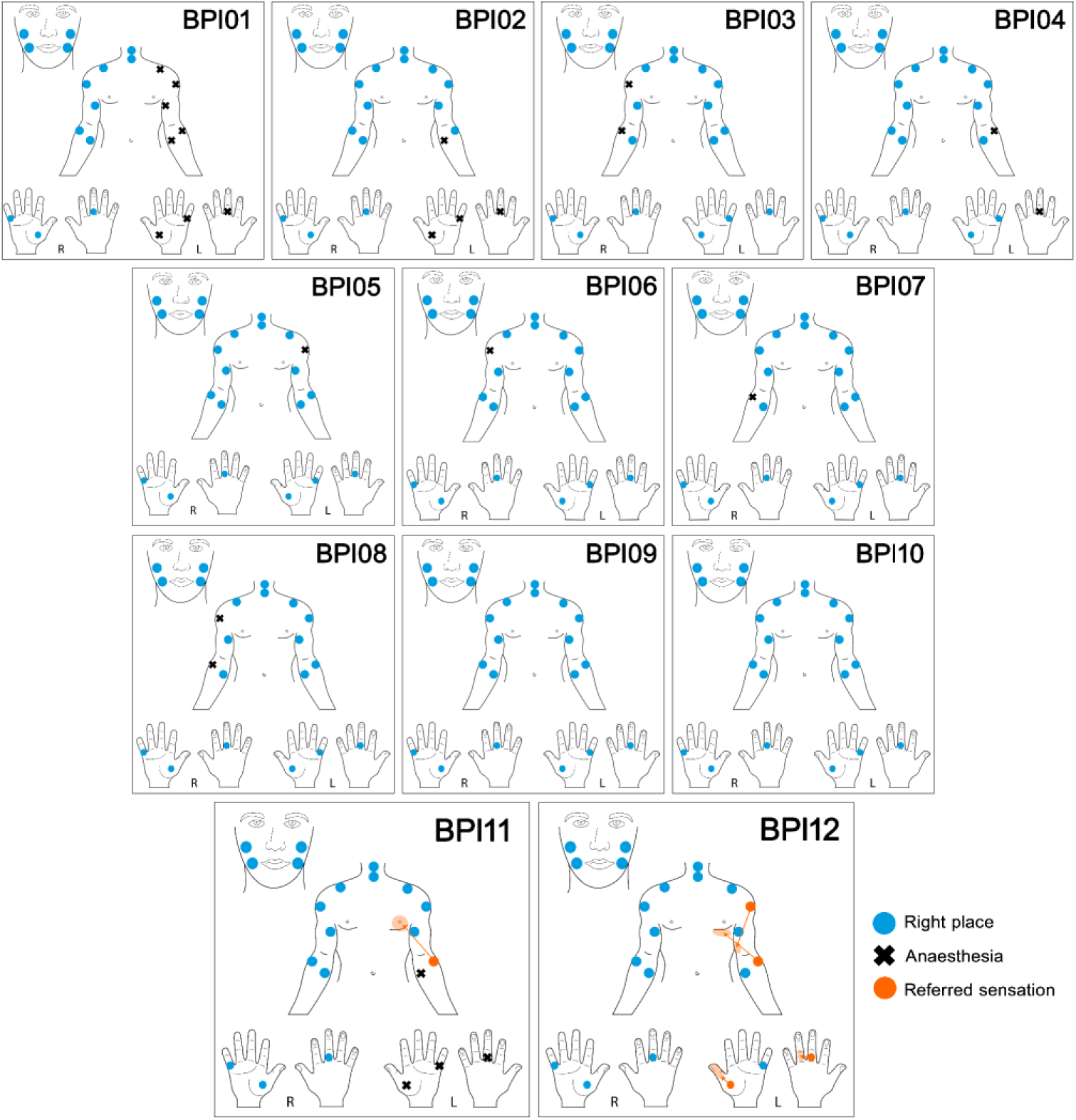
Screening of referred sensations from BPI01 to BPI12. The blue circles represent the points where sensations were evoked in the correct place. Black crosses represent the points of anaesthesia. Orange circles represent the points that evoked referred sensations (indicated by the arrows).

### Experiment 2: Mapping of the forearm referred sensation (RS)

Subjects who reported RS (BPI11, BPI12, and BPI13) will be described separately below. All subjects had a motorcycle accident that led to complete left BPI. Information about the participants, type of surgery, time between injury and surgery, and time between injury and sensory assessment are shown in Table 1.

#### BPI 11

The sensory mapping of the patient BPI11 forearm can be seen in Figure 3. In total, 46 points were stimulated in the upper limb (Fig 3A), resulting in a grid of sensory responses represented in Figure 3B. From this total, 21 were considered anaesthesia points (crosses in Fig. 3A, B). One point evoked a spatially correct sensation (open circle in Fig. 3A, B), 9 points evoked RS in the upper limb (grey squares in Fig. 3A, B) and 15 points evoked RS in the chest (black squares in Fig. 3A, B). These 24 RS points were enumerated, so the precise place of the RS can be seen in Fig 3C. The presence, topological distribution and extent of the RS mapped in the chest and forearm can be appreciated in figure 3D. RS evoked when stimulating forearm points were spread all over the left chest, without any identified type of topographical organisation. Some stimulated points led to unpleasant sensations (pricking sensation - red stars in Fig. 3D). The sensations evoked in the forearm (1-9 grey squares in Fig. 3B) were always identified in the same spot (“a” area on the forearm, Fig. 3D).

**Figure 3.**
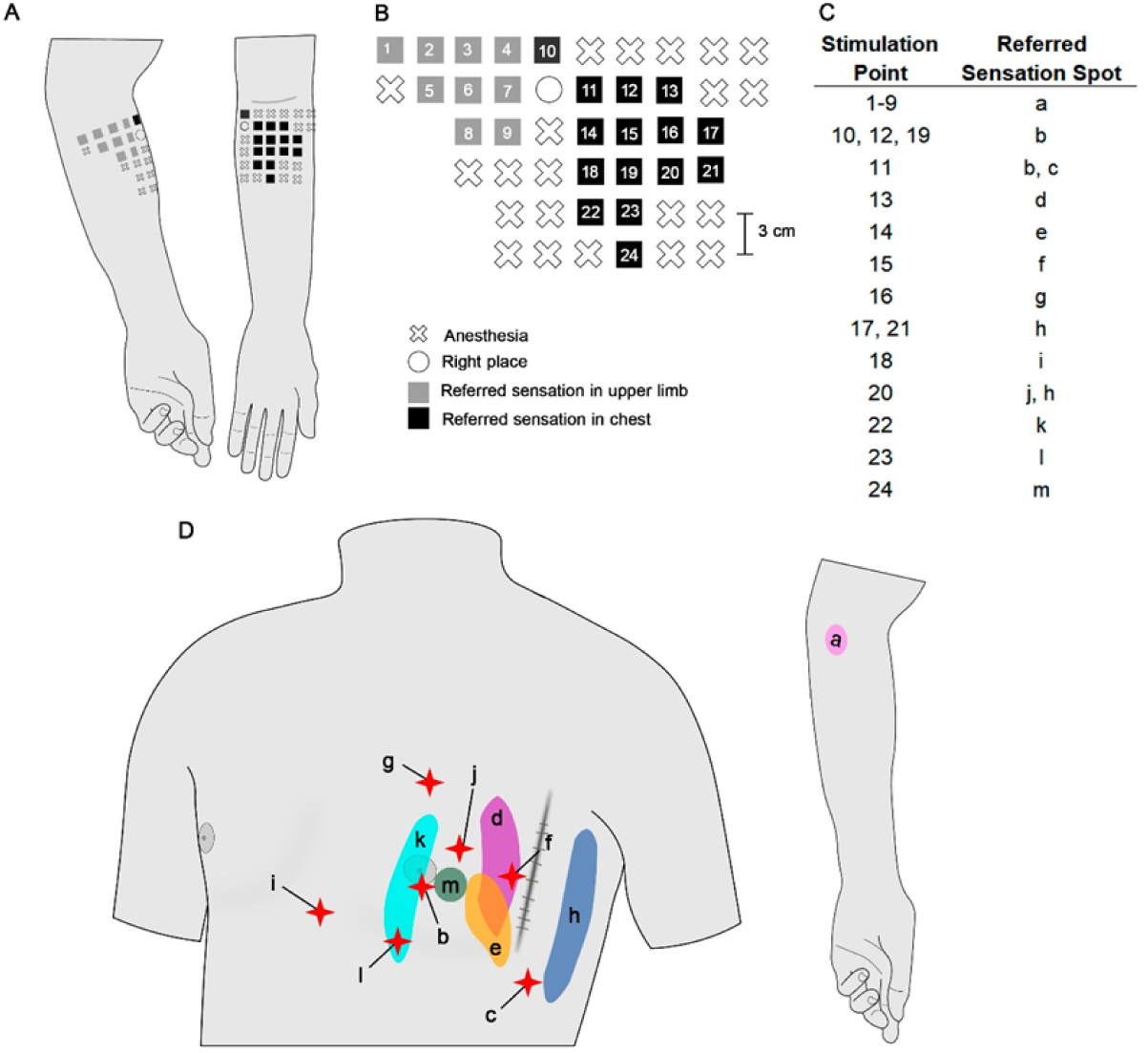
Referred sensation mapping of BPI11. **(A)** Representation of the stimulated points grid of the forearm. **(B)** Stimulated points grid - each point is represented according to the participant’s report. **(C)** Table listing the points that evoked RS on the forearm and the sites of the RS, which can be seen in D. **(D)** Representation of the RS sites. Red stars represent a pricking sensation.

#### BPI 12

The sensory mapping of the forearm of patient BPI12 can be observed in Figure 4. In total, 81 points were stimulated, forming the grid represented in Fig 4A and B. Six were considered as anaesthesia points (crosses in Fig. 4A, B), and 48 points evoked correctly placed sensations (open circles in Fig. 4A, B). Thirteen points evoked both correctly placed sensations and RS in the chest (black circles in Fig. 4A, B), three points evoked RS in the chest (black squares in Fig. 4A, B), six points evoked RS in the upper limb (grey squares at Fig. 4A, B) and five points evoked RS both in the chest and the upper limb (grey squares with black border at Fig. 4A, B). The RS points were enumerated, so the precise place of the RS can be observed in Figure 4C and D.

**Figure 4.**
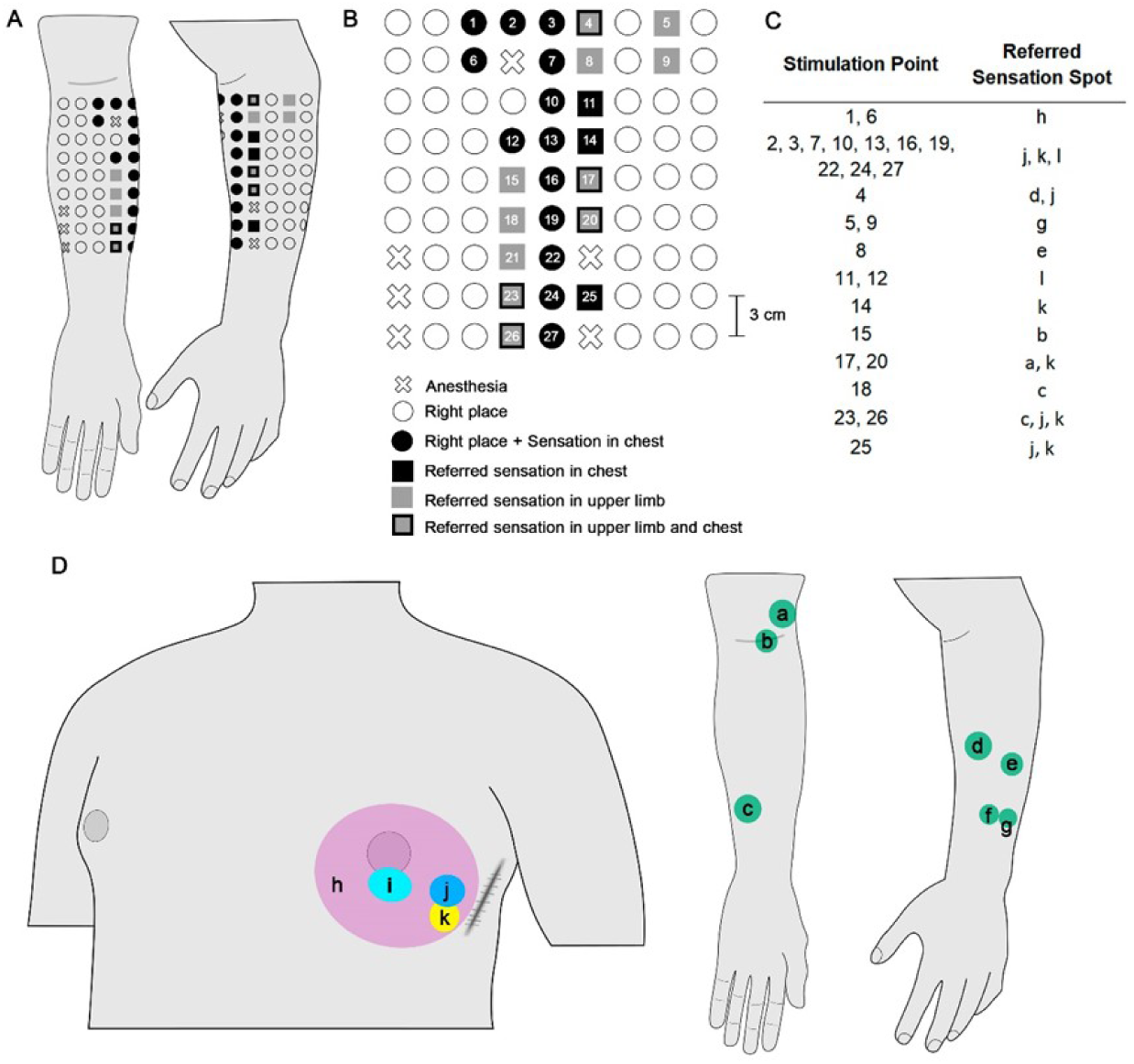
Referred sensation mapping of BPI12. **(A)** Representation of the stimulated points grid of the forearm. **(B)** Stimulated point grid - each point is represented according to the participant’s report. **(C)** Table listing the points that evoked RS on the forearm and the location of the RS, which can be seen in D. **(D)** Representation of the RS sites.

Unlike subject BPI11, BPI12 showed a concentration of RS in the chest while the sensations evoked in the forearm were spread. This participant also presented grid points (grey squares with black borders, Fig. 4B) in which the stimulation was felt both on the chest and the forearm (Fig. 4C, D).

#### BPI 13

BPI13 was assessed three times over a time span of 6.3 years. The interval between injury and each assessment was 165, 844, and 2328 days (5.5 months, 2.3 years, and 6.3 years, respectively). In its first assessment, BPI13 did not report any RS in the chest when stimulated in his left forearm. Furthermore, the musculocutaneous point of exclusive innervation was devoid of any sensation. The mapping performed in the second assessment is shown in Figure 5A-D. The grid had 42 stimulated points, where 36 were considered anaesthesia points (crosses in Fig. 5A, B). Six points evoked RS in the chest (black squares at Fig. 5A, B), always at the same place (Fig. 5C, D).

**Figure 5.**
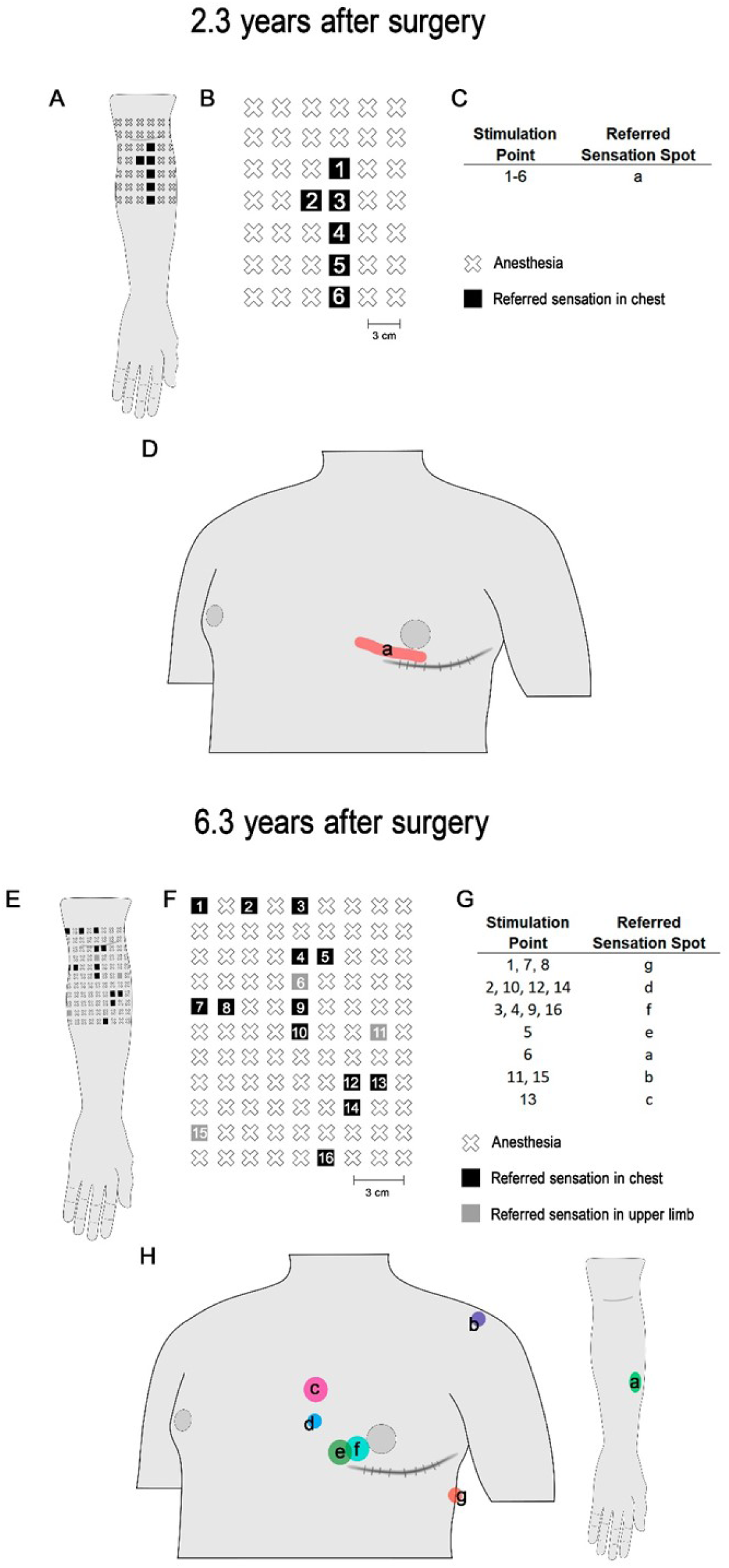
Second and third referred sensation mapping of BPI13. **(A)** and **(E)** representation of the stimulated point grid of the forearm. **(B)** and **(F)** Stimulated point grid - each point is represented according to the patient’s report. **(C)** and **(G)** Table listing the points that evoked RS on the forearm and the sites of the RS, which can be seen in D and H. **(D)** and **(H)** Representation of the RS sites.

Finally, mapping was performed again in the third assessment (Fig. 5E-H). At this time, from a total of 99 points stimulated, 83 were considered anaesthetised points (crosses at Fig. 5E, F), 13 points evoked RS in the chest (black squares at Fig. 5E, F) and three points evoked RS in the forearm (grey squares at Fig. 5G, H).

Notably, when comparing the two mappings, it is possible to see a scattering of RS over the course of time. At the first mapping (2.3 years after surgery), stimulated points that evoked RS were close to each other, aligned longitudinally along the forearm, and all of them evoked RS at the same site on the chest. Otherwise, after 4 years, RS were evoked by stimulating points spread across the forearm. Furthermore, the RS evoked at the chest were diffuse, and there was also an RS in the forearm.

## Discussion

In this study, a systematic and standardised assessment was carried out to screen referred sensation (RS) in 13 participants with TBPI. Each participant (except one) underwent at least one out of four types of nerve transfer. RS was only identified in the three patients who underwent ICNs-MCN transfer. For each patient, the RS systematic mapping revealed a unique distribution in the forearm and the chest, without any clear topographic organisation. Pricking and simultaneous chest/forearm RS were also reported. Longitudinal assessment in one of the tested participants showed a scattered expansion of the RS throughout time. This is the first study to systematically evaluate this phenomenon in patients with TBPI.

These results are in accordance with previous reports of the presence of RS after ICNs-MCN transfer aiming at the biceps motor recovery ^22,30,31,38–41^. It is important to highlight that the referred sensation screening included clear and systematic instructions to participants, aiming to avoid inductions of false RS as already demonstrated ^34^. Confirming the results found in our sample, and to the best of our knowledge, no RS was ever reported after an Oberlińs nerve transfer nor after a radial to axillary nerve transfer, or after an accessory to suprascapular nerve transfer. One possible explanation for the development of RS after ICNs-MCN transfer is the large amount of sensory fibers present in the donor nerve ^42^. Even if there is an attempt to isolate the motor component of the intercostal nerve, some sensory fibers may be transferred and cause sensory reinnervation of the forearm, leading to RS. In contrast, the other techniques use mainly motor nerves or motor branches as donor and receptor nerves ^43–45^, probably restricting any sensory nerve transfer.

Since the aim of nerve transfer surgeries in TBPI patients is motor recovery, the sensory outcomes of this technique are commonly described as a secondary effect or an adverse result. Nevertheless, sensory reinnervation has been investigated after peripheral nerve injuries of the upper extremity, primarily aiming at restoring the hand’s protective sensation ^46,47^. In this technique, the ulnar nerve and the lateral cord of the median nerve are used as main receptor nerves to restore the sensitivity of the hand and fingers ^47^. Likewise, the sensory branch of the MCN is used to reinnervate the skin of the lateral forearm ^48,49^. Some of the possible nerve donors for sensory reinnervation are the intercostal nerve ^40,48,50–54^ and the contralateral C7 root ^40,49,55^. Thus, also in these cases, as a consequence of an intercostal nerve transfer, the stimulation of the hand or the lateral forelimb territory often leads to the perception of the sensation at the chest ^39,50,56,57^. When RS arises after nerve transfer and sensation is evoked at the original donor nerve territory, it is referred to as “right way” ^4^.

One interesting finding is that participant BPI11, who was evaluated at the earliest time after surgery, reported feeling pricking RS. This is in line with data that have been reported since the beginning of the 20th century, with Rivers and Head (1908) (apud ^58^ who observed that pain and heat sensations are the first to return after a nerve injury and following suture, in their case of the median nerve. These types of sensations, as well as itching and ‘tingling’ (assessed by the Tinel’s test), are carried mainly by unmyelinated fibres, possibly c-fibres, which are the first to regenerate ^58–60^.

In our sample, “Right way” RS were typically scattered, without any indication of topographic distribution in the chest and forearm. Interestingly, the longitudinal mapping (up to 6.3 years after surgery) of RS in one patient (BPI13) revealed a progression in the RS distribution, from a single RS localised in the chest, progressing towards a higher number of RS with spread distribution, suggesting a lack of direction in the regeneration of the peripheral nerve components. These results agree with previous reports of the absence of any topographic specificity regarding sensory nerve regeneration ^61–63^. After nerve repair, nerve fibres can also grow misdirected, achieving the wrong/non-intended skin regions. When it happens, the touch in the reinnervated area is perceived in the original skin territory of the growing nerve, or the donor’s nerve, in the case of nerve transfer ^59,64^.

The longitudinal mapping also revealed the temporal pattern of RS appearance and persistence: RS was absent during the first 5.5 months after surgery, being reported after 2.3 years, and persisting over a period of 6.3 years. These findings are in line with reports describing the RS onset at around 12 months after a nerve transfer ^30,31^, being found even after 6 years post-surgery ^22,41^. For instance, Nagano et al. (1989) found that most of the 176 patients that underwent the ICTs-MCN transfer presented the “right way” referred sensation. Interestingly, 6 years after surgery, the sensation in the musculocutaneous territory gradually began to appear at some degree ^41^. The only three cases that showed complete switching of the sensibility from the chest to the upper limb were under 20 years of age (19, 11 and 11 years old), operated at the ages of 3, 4, and 8 years, respectively ^41^. The RS persistence after sensory nerve transfer in adult TBPI patients might indicate a peculiar reorganisation in the topographical representations in the primary somatosensory cortex (S1).

Curiously, such reorganisation has been employed as a tool in amputees to provide useful sensory feedback from a prosthetic device ^35,36,65^. The “target reinnervation” consists of transferring residual arm nerves to chest muscles or to the stump in order to enable the growth of sensory afferents to reinnervate the chest or the stump skin. When the sensory reinnervation takes place, the tactile stimulation in the reinnervated area evokes the sensation on their missing limb ^35,36,65^. The persistence of this sensation has been reported in patients up to 16 years after the “target reinnervation” procedure ^65^.

The rearrangement in S1 is expected after peripheral injury and nerve transfer ^16,66^. Electrophysiological recording in primates’ brain mapping after restricted deafferentation showed an invasion of adjacent representations on the deprived cortex from remnant inputs in S1 ^67^. However, when the sensory loss is extensive, part of S1 may remain unresponsive ^68^. Furthermore, when nerve regeneration occurs, previously well-defined cortical representations are transformed into scattered discontinuous representations due to the misdirection of axonal growth ^66^. A very early report in rats ^69^, after having their nerves mediating cutaneous and deep sensibility crossed from the left hind foot to the right, persisted in withdrawing the unstimulated leg (e.g. right leg) when the sole of the foot (left leg) was stimulated with an electric shock. The author argued that the persistence of the original reflex patterns without central adjustments after the crossed reinnervation reveals an extreme stability in the reflex circuits from the sensory to motor side ^69^. More recently, Chen et al. (2007) performed a study with 12 infants with birth injury or TBPI who underwent C7 contralateral root transfer (crossed reinnervation) at the age of 6 to 93 months. In a follow-up of 21 to 63 months after surgery, all patients presented “little” or “substantial” synchronous motion and sensibility at the donor limb, which means, movement or sensation at the donor limb which accompanied function of the nerve repaired on the affected side. Together, these findings reinforce a constrained S1 reorganisation capacity, which could justify the RS persistence over the longitudinal mapping after nerve transfer.

Conversely, after an ICNs-MCN transfer, biceps function recovery goes along with topographic reorganisation in the primary motor cortex (M1) ^70–72^. Investigating longitudinal changes in M1 of these patients, Mano et al. (1995) found that up to six months after the surgery, the biceps representation in M1 was spatially closer to the representation of the trunk. Also, biceps contraction was always associated with respiratory movements ^70^. Then, one to two years after ICNs-MCN nerve transfer, as the patients acquired the ability to flex the elbow in dissociation with respiratory movements, and the biceps representation became independent and more lateral to that of the intercostal r in M1 (see also ^71^. Corroborating these findings, employing a fMRI protocol, Malessy et al. (2003) showed that, after at least 4 years of surgery, the centre of gravity of the reinnervated biceps representation appeared more laterally in M1 than that of the intercostal muscle representation. Unfortunately, these studies did not mention the sensory component outcomes. The assessment of cortical reorganisation of adult TBPI patients through fMRI after C7 contralateral root transfer to the biceps muscle showed that, around 13 to 36 months after surgery, elbow flexion with the neurotized arm was associated with bilateral M1 activation ^73^. Thus, it can be considered that the cortex ipsilateral to the lesion, even 3 years after surgery, remains responsible for activating the donor nerve, participating in the elaboration of movement jointly with the contralateral cortex ^73^.

Therefore, it appears that for successful nerve transfer, the donor and recipient nerves must have closer cortical sensorimotor representations and also display similar functional behaviour. Coordination between arm and trunk during reaching or pointing movements or anticipatory trunk postural adjustments during upper limb movements have already been described ^74,75^. However, there seems to be little sensory synergy between these parts; that is, the lateral forearm skin (musculocutaneous sensory innervation) and the chest skin (intercostal sensory innervation) do not have a clear sensory interaction. This disconnected sensory function between the chest and the lateral forearm can make it difficult to reestablish the localization perception after ICN-MCN transfer; in other words, the patient continually feels a lateral forearm stimulation as a chest sensation.

Among the limitations of the present study, we acknowledge the small sample size of patients. Furthermore, more detailed surgical information might be useful to correlate with the onset and distribution of RS in the chest in future studies.

## Conclusions

Among the reported types of nerve transfer, only the patients who underwent ICN-MCN transfer presented RS. A possible explanation for this phenomenon is that the great distance between the representations of the donor and receptor nerves at the primary somatosensory cortex (S1) could constrain plastic reorganisation and thus limit the disentangling of the forearm and chest sensations. This information may be useful to guide future surgical and rehabilitation protocols concerning peripheral nerve transfers.

## Data Availability

All data produced in the present study are available upon reasonable request to the authors

## Author Contributions

A.C.S, F.S.E, C.D.V. and B.L.R conceived and planned the experiments. A.C.S. and B.L.R. carried out the experiments. A.C.S., C.D.V. and B.L.R contributed to the interpretation of the results. A.C.S. and B.L.R. took the lead in writing the manuscript. All authors provided critical feedback and helped shape the research, analysis and manuscript.

